# Long term anti-SARS-CoV-2 antibody kinetics and correlate of protection against Omicron BA.1/BA.2 infection

**DOI:** 10.1101/2022.12.13.22283400

**Authors:** Javier Perez-Saez, María-Eugenia Zaballa, Julien Lamour, Sabine Yerly, Richard Dubos, Delphine Courvoisier, Jennifer Villers, Jean-François Balavoine, Didier Pittet, Omar Kherad, Nicolas Vuilleumier, Laurent Kaiser, Idris Guessous, Silvia Stringhini, Andrew S. Azman, the Specchio-COVID19 study group

## Abstract

Binding antibody levels against SARS-CoV-2 have shown to be correlates of protection against infection with pre-Omicron lineages. This has been challenged by the emergence of immune-evasive variants, notably the Omicron sublineages, in an evolving immune landscape with high levels of cumulative incidence and vaccination coverage. This in turn limits the use of commercially available high-throughput methods to quantify binding antibodies as a tool to monitor protection at the population-level. In this work, we leverage repeated serological measurements between April 2020 and December 2021 on 1’083 participants of a population-based cohort in Geneva, Switzerland, to evaluate anti-Spike RBD antibody levels as a correlate of protection against Omicron BA.1/BA.2 infections during the December 2021-March 2022 epidemic wave. We do so by first modeling antibody dynamics in time with kinetic models. We then use these models to predict antibody trajectories into the time period where Omicron BA.1/BA.2 were the predominant circulating sub-lineages and use survival analyses to compare the hazard of having a positive SARS-CoV-2 test by antibody level, vaccination status and infection history. We find that antibody kinetics in our sample are mainly determined by infection and vaccination history, and to a lesser extent by demographics. After controlling for age and previous infections (based on anti-nucleocapsid serology), survival analyses reveal a significant reduction in the hazard of having a documented positive SARS-CoV-2 infection during the Omicron BA.1/BA.2 wave with increasing antibody levels, reaching up to a three-fold reduction for anti-S antibody levels above 800 IU/mL (HR 0.30, 95% CI 0.22-0.41). However, we did not detect a reduction in hazard among uninfected participants. Taken together these results indicate that anti-Spike RBD antibody levels, as quantified by the immunoassay used in this study, are an indirect correlate of protection against Omicron BA.1/BA.2 for individuals with a history of previous SARS-CoV-2 infection. Despite the uncertainty in what SARS-COV-2 variant will come next, these results provide reassuring insights into the continued interpretation of SARS-CoV-2 binding antibody measurements as an independent marker of protection at both the individual and population levels.

## Introduction

While by mid-2022 a large fraction of the global population had developed anti-SARS-CoV-2 binding antibodies through infection and/or vaccination (Bergeri et al., 2022; Zaballa et al., 2022), it remains unclear whether seroprevalence results translate into prevalence of effective protection against infection (Theel et al., 2020). Neutralizing antibodies may provide a reliable correlate of protection against both infection and severe disease (Earle et al., 2021; Gilbert et al., 2022; Khoury et al., 2021; Krammer, 2021). Neutralization assays are, however, labor-intensive and challenging to use at a large scale, despite advances in high-throughput surrogate assays (Fenwick et al., 2021).

Binding antibody measurements have been found to correlate with neutralization capacity against the ancestral SARS-CoV-2 strain at different degrees depending on time post infection/vaccination, and on immuno-assay (Earle et al., 2021; Goldblatt et al., 2022; L’Huillier et al., 2021). Evidence for their more general use as correlate of protection is mounting both from population-level (Earle et al., 2021; Goldblatt et al., 2022), as well as individual-level studies in the context of vaccine trials (Dimeglio et al., 2022b; Feng et al., 2021; Gilbert et al., 2022; Wei et al., 2021). These studies suggest that higher antibody measurements after infection and/or vaccination tend to reduce subsequent infection risk (Osman et al., 2021). However, most of these studies focused on the ancestral SARS-CoV-2 strain and it is yet to be clarified whether these results robustly extend to the Omicron subvariants (Hertz et al., 2022; Zar et al., 2022).

The evaluation of binding antibody levels as correlates or protection is challenged by the constant evolution of the anti-SARS-CoV-2 immune landscape through vaccination and successive epidemic waves driven by different virus variants. Longitudinal antibody studies up to 14 months follow-up have shown that antibody levels change with time since infection and/or vaccination across individuals, and depending on the immuno-assays used for detection (Eyran et al., 2022; Gallais et al., 2021; Peluso et al., 2021; Perez-Saez et al., 2021). Characterization of long-term antibody kinetics provides an opportunity for leveraging serological cohort studies to complement vaccine trials in evaluating binding antibody levels as correlate of protection against future infections. By relying on binding antibody immunoassays that are simple, standardized and widely used worldwide, the results of these studies have the potential to be generalized to other settings despite their functional limitation. These cohort studies might therefore contribute by assessing the extension of results to different commercially-available immunoassays and a wider range of infection/vaccination histories, in particular to non-vaccinated individuals.

In this study, we aim to evaluate the use of a commercially available immunoassay as a correlate of protection in the Omicron era. We do so by leveraging repeated serological measurements and reported infections on a population-based longitudinal cohort followed for up to 20 months in the state of Geneva, Switzerland. We first characterize antibody dynamics during the longitudinal serology period (April 2020 to December 2021) using kinetic models fit to observed antibody measurements. We then project each individual’s antibody trajectories into the Omicron exposure period (December 2021 to March 2022) to explore the relationship between projected antibody levels and having a SARS-CoV-2 positive test.

## Materials and methods

### Study design

This study uses data from the population-based Specchio-COVID19 cohort, composed of adult participants recruited through serological surveys (Baysson et al., 2022; Stringhini et al., 2021a, 2021b, 2020). Following their baseline serology, participants in this cohort are regularly invited to complete online questionnaires, where they report SARS-CoV-2 test results, disease severity and vaccination status, and can be proposed one or several serological tests during the follow-up. Each participant coming for a follow-up serology provided a venous blood sample and filled in a short paper questionnaire on site to update/complete their information on infection and vaccination statuses.

In this study, our main analysis consisted of two steps. Firstly, we analyzed antibody trajectories during the longitudinal serology period, when serological testing follow-up was conducted (April 6th 2020, to December 17^th^ 2021). Secondly, we evaluated correlates of protection against infection during the “exposure period”, using information on SARS-CoV-2 infections from the surge of the Omicron BA.1 subvariant in the state of Geneva until the end of the study period (December 25^th^ 2021 to March 20^th^ 2022) (Figure 1). In this period there were no specific quarantine and isolation measures following SARS-CoV-2 infection nor other specific recommendations in Geneva, which may have contributed to low virological testing rates. During the exposure period Omicron BA.1 and BA.2 subvariants comprised nearly all infections. From all participants of the Specchio-COVID19 cohort, in this analysis, we only included those having at least two positive serologies and for whom we had complete vaccination information by March 20^th^ 2022 (Supplementary Material Figure S1). During the study period, the only available COVID-19 vaccines in Switzerland were the mRNA-BNT162b2/Comirnaty from Pfizer/BioNTech (since December 2020), mRNA-1273 from Moderna/US NIAID (since January 2021), and the Janssen Ad26.COV2.S COVID-19 vaccine (since October 2021).

**Figure 1.**
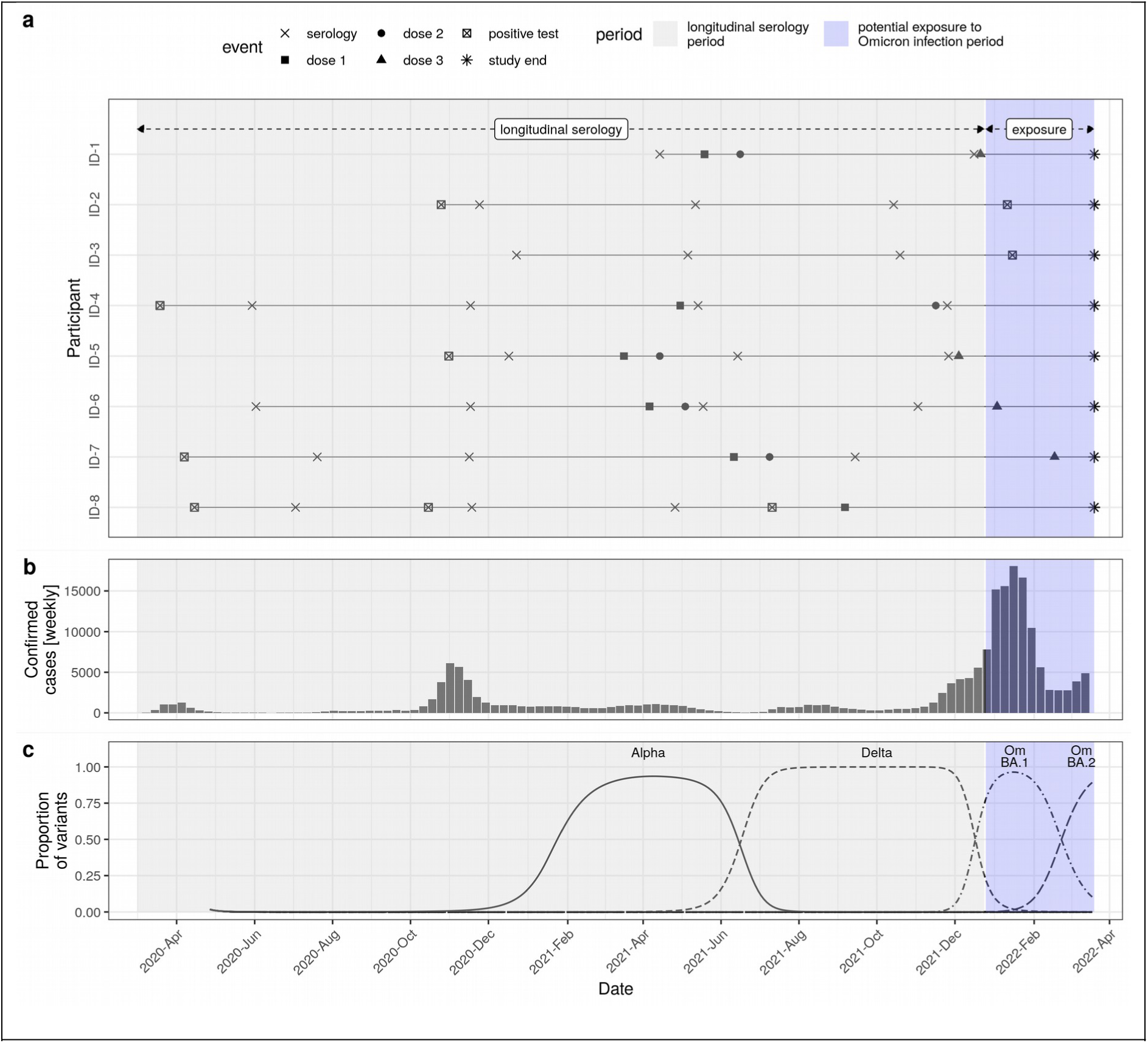
Study context. a) Study phases and eight examples participant-level data. b) Weekly confirmed SARS-CoV-2 cases in the state of Geneva (available from: https://infocovid.smc.unige.ch). c) Proportion of SARS-CoV-2 variants in sequences sampled in Western Switzerland estimated through multinomial spline regression of publicly available weekly sequence data from the Covariants project (https://github.com/hodcroftlab/covariants/), following analysis from https://www.hug.ch/laboratoire-virologie/surveillance-variants-sars-cov-2-geneve-national (report May 2022).

This study was approved by the Geneva Cantonal Commission for Research Ethics (CCER project number 2020-00881) and written informed consent was obtained from all participants.

### Immunoassays

For this study, we used the quantitative Elecsys anti-SARS-CoV-2 RBD immunoassay, which measures total antibodies (IgG/A/M) against the receptor binding domain of the virus spike (S) protein (#09 289 275 190, Roche-S, Roche Diagnostics, Rotkreuz, Switzerland). Seropositivity was defined using the cut-off provided by the manufacturer of ≥0.8 U/mL. Output test values were transformed to WHO international standard units by multiplying by a factor of 1.184. We calculated the intra-lot coefficient of variation (CV) for each batch of our internal positive control serum and the maximum CV (7.3%) was used to define uncertainty in serological measurements in the kinetic model described below (Supplementary Material Section S2). To identify past infections in vaccinated participants, we also measured total levels of antibodies binding the nucleocapsid (N) protein using the semiquantitative Elecsys anti-SARS-CoV-2 N immunoassay (#09 203 079 190, Roche-N). The three vaccines available in Switzerland during the study period elicit a response exclusively to the Spike protein of SARS-CoV-2, as opposed to infection, typically eliciting a response to both the N and S virus proteins. Although not the focus of the main analysis, we also present anti-N antibody trajectories in the Supplementary Material (Supplementary Material Section S3).

### SARS-CoV-2 virological tests data

For the correlate of protection analysis, results of PCR and antigenic tests were extracted from the ARGOS database up to March 20^th^ 2022. The ARGOS database consists of a general register of COVID-19 diagnostic tests performed in the state of Geneva since February 2020 and is maintained by the state directorate for health (Genecand et al., 2021). Data on test results from ARGOS were supplemented with additional information on COVID-19 diagnostic tests (PCR or antigen-based rapid diagnostic tests [RDTs] including self-tests) as self-reported by the participants through regular questionnaires.

### Statistical analyses

#### Antibody trajectories analysis

In the first step of the analysis (“longitudinal serology” period in Figure 1), we characterized antibody dynamics by fitting the observed antibody trajectories to bi-phasic kinetic models (Pelleau et al., 2021). These models assume an initial post-infection/vaccination antibody boost (increase in antibody levels at a given time post-exposure) followed by initially fast then slower exponential decay. We here expanded these models to account for multiple boosting events due to infection and/or vaccination. The size of antibody boosts and decay in time are determined by age, sex and boosting history (the sequence of infections and vaccine doses). We further accounted for observed individual-level variability in anti-SARS-CoV-2 antibody kinetics. Inference was performed in a Bayesian hierarchical framework incorporating uncertainty of the timing of infection events in the absence of information on COVID-19 diagnostic tests. Model details are given in the Supplementary Material (Section S4). Stan model code and minimal testing datasets are available at https://github.com/UEP-HUG/serosuivi_2021_public.

#### Survival analysis

In the second step of the analysis, we evaluated binding antibody levels as a correlate of protection against Omicron BA.1/BA.2 infections during the “exposure period” (Figure 1) using survival analysis methods. The aim was to infer the effect of being above different thresholds of binding antibody levels (as measured by the Roche-S immunoassay) on the hazard of confirmed SARS-CoV-2 infection during the exposure period. We focused on the exposure period from when Omicron BA.1 became dominant (more than 80% of samples on Dec 25^th^ 2021) up to the latest date for which we had access to the state registry of SARS-CoV-2 test results (March 20^th^ 2022). By this date Omicron BA.2 had replaced Omicron BA.1 in Western Switzerland (90% vs. 10% of typed samples, Figure 1). We excluded participants with either (a) no serology information after April 1^st^ 2021, or (b) uncertain infection status during the Omicron exposure period due to missing data (i.e. missing positive test result and missing self-reported absence of positive tests, details in Supplementary Material Section S1). We used Cox proportional hazards model, controlling for age and previous infection status based on our assumptions of the relationship between variables (Supplementary Material Section S5). Given that we used modeled antibody levels during the exposure period based on available serological prior measurements, we excluded participants for whom a boosting event (infection and/or vaccination) occurred between the last serological measurement and the start of the exposure period if the modeled antibody level was below the threshold of interest (Supplementary Material Section S1). Potential informative censoring due to vaccination and/or infection during the Omicron exposure period was adjusted for through inverse probability weighting as implemented in the ipw R package (Wal and Geskus, 2011).

## Results

The cohort included in this study was composed of 1’083 adult participants, 55% of whom were female and 91% were younger than 65 years (Table 1). Participants in the cohort had few comorbidities and no immunosuppressive diseases in general. Among participants, 91% had a history of SARS-CoV-2 infection (based on positive tests or anti-N serology as defined in the Supplementary Material Section S1) and 58% were unvaccinated at the time of their most recent serology (range of last serology dates November 16^th^ 2020 – December 17^th^ 2021, median June 21^st^ 2021). Around half of participants with a history of infection based on a positive anti-N serology did not have any diagnostic screening tests during the actue phase of their infection (PCR or RDT) (58%). Most participants in the cohort had only two positive serological tests (56%), and 10% had 4 or more seropositive samples. All event data is presented in Supplementary Figure S2.

**Table 1.** Participant characteristics. Characteristics are given for the five encountered infection and vaccination history.

| Characteristic | N = 1'083<sup>1</sup> |
| --- | --- |
| **Sex** |  |
| female | 590 (55%) |
| male | 493 (45%) |
| **Age (years)** |  |
| 18-64 | 988 (91%) |
| 65+ | 95 (9%) |
| **Infection/vaccination status at time of most recent serological test** |  |
| infected only | 634 (58%) |
| vaccinated only | 98 (9%) |
| infected prior to vaccination | 310 (29%) |
| infected after vaccination | 11 (1%) |
| infected prior to vaccination and re-infected after vaccination | 30 (3%) |
| **Virological confirmation among participants with history of infection (N=985)** |  |
| Virologically confirmed infection (self-reported or from ARGOS registry) | 567 (58%) |
| No virological confirmation (anti-N positive serology) | 418 (42%) |
| **Vaccine type among vaccinated participants at time of last serological status (N=449)** |  |
| mRNA-1273 (Moderna/US NIAID) | 247 (55%) |
| mRNA-BNT162b2/Comirnaty (Pfizer/BioNTech) | 202 (45%) |
| **# Positive Roche-S serological samples per participant within this study** |  |
| 2 | 603 (55%) |
| 3 | 374 (35%) |
| 4 | 104 (10%) |
| 5 | 1 (<0.1%) |
| <sup>1</sup>n (%) |  |

### Antibody trajectories during the longitudinal serology period

Serological samples were collected from 1’083 participants between April 2020 and December 2021, with follow-up times between first and last visits of up to 20 months following infection and up to 8 months following vaccination (Figure 2a). Over this longitudinal serology period, we did not observe anti-S-based seroreversion for any participants (Figure 2b). Antibody levels following vaccination were distributed in the upper range of the immunoassay’s dynamic range (Figure 2b, Supplementary Figure S3), with more than one in three samples collected at least 14 days after participant’s latest vaccine dose having values above the upper quantification limit of the test (33%, Figure 2d). Anti-N antibody trajectories are shown in Supplementary Figure S4.

**Figure 2.**
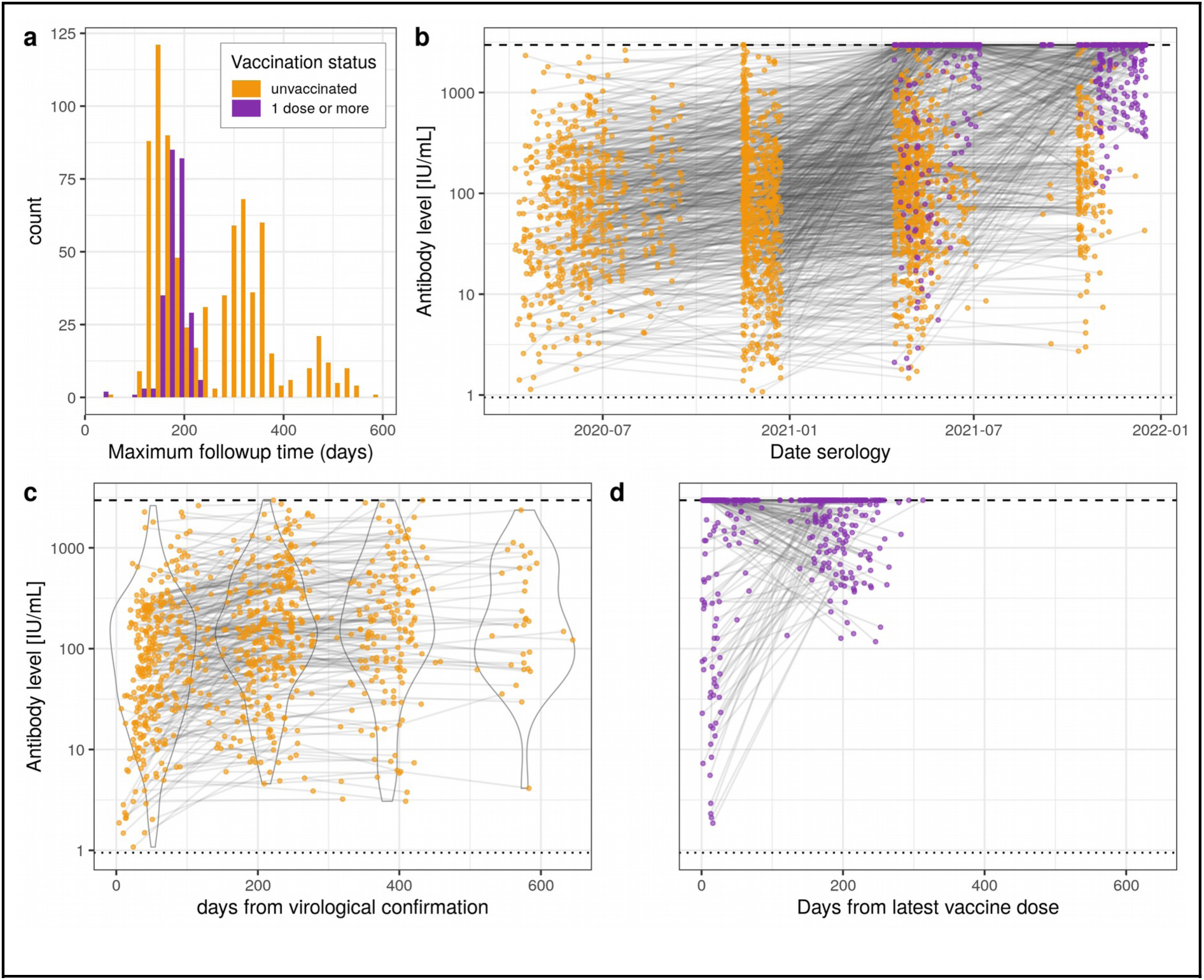
Anti-S binding antibodies level trajectories. a) Follow-up time distribution (time from participant’s first to last serology) for samples collected prior (n = 778) and post (n = 246) vaccination when at least two positive samples were available. Note that participants may have multiple samples prior and post vaccination and may therefore appear in both categories. b) Trajectories for all participants (n = 1’083) by serological sampling date and according to vaccination status. c) Trajectories of pre-vaccination samples by time from virological confirmation when available (n = 442), along with violin plots of antibody levels in discrete arbitrary categories of time post confirmation (0-149, 150-249, 250-449, 450+ days). d) Trajectories post-vaccination by time from latest dose (n = 246). Dashed and dotted lines in panels b, c and d indicate upper quantification limit (2500 U/mL, equivalent to 2960 IU/mL) and threshold for positivity (0.8 U/mL, equivalent to 0.95 IU/mL) of the test, respectively.

To investigate how infection and vaccination history affects antibody levels, we fit kinetic models to individual-level antibody trajectories. Mean antibody rises were similar among age classes. Rises in anti-S binding antibody levels depended markedly on both infection and vaccination history (Figure 3a, parameter estimates in Supplementary Table S2). The weakest estimated anti-S boost were the ones following infection in unvaccinated individuals, while the strongest boost was estimated following the first vaccine dose in previously infected individuals. Among vaccinated and infected individuals the estimated anti-S boost parameter decreased with the number of vaccine doses. Among uninfected individuals the largest increase in anti-S levels occurred after the second vaccine dose, with similar levels for the first and third doses. Mean antibody half-lives showed less variation among boosting events, ranging from 50 days (95% CrI 30-100) following the second vaccine dose in uninfected 18-64y individuals to 510 days (140-1360) in 65+ individuals with two infections and one vaccine dose (Figure 3b). Estimated antibody half-lives were similar across individuals both infected and vaccinated, regardless of the number of vaccine doses received. In turn, antibodies decayed faster in uninfected individuals following the second dose, as opposed to antibodies mounted with the first and third doses. Both boost level and antibody decay rates had considerable although uncertain individual-level heterogeneity with coefficients of variation of 5.1 (95% CrI 0.2-50.0) and 22.1 (0.3-65.5), respectively. These kinetic parameter estimates along with inference on individual-level variability allowed us to model antibody trajectories for each participant with strong agreement with available serological measurements (Figure 3c, Supplementary Figure S5).

**Figure 3.**
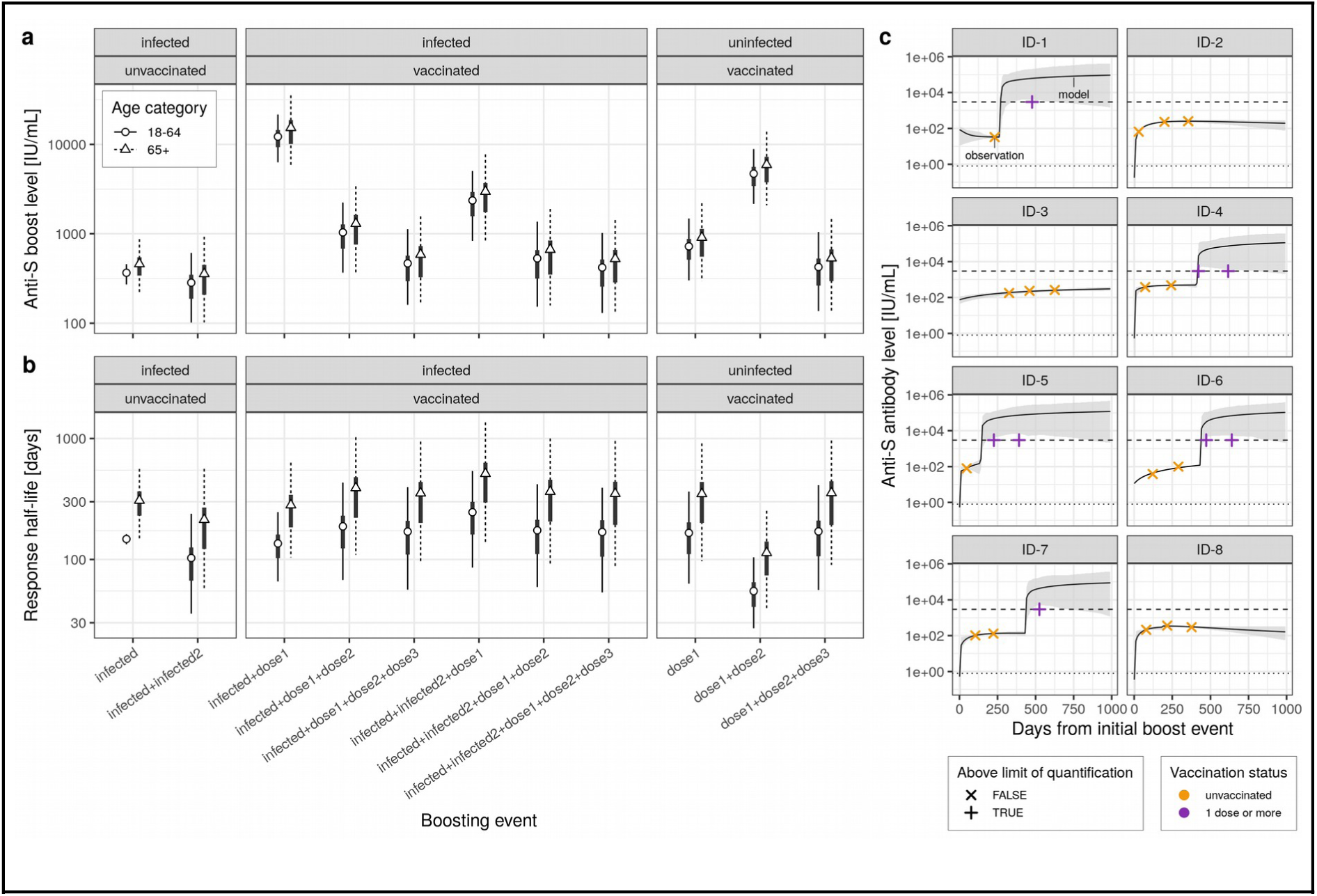
Antibody dynamics inference. a) Inferred mean antibody level boosts following infection and/or vaccination by age category and infection/vaccination history (dots: mean, thick lines: 50% CrI, thin lines: 95% CrI). “Dose1/2/3” denotes the vaccine dose, and “infected1/2” denotes the infection (first or second infection). Note that the order of boosting events is not taken into account in the model, and that boosting events are considered to be additive. b) Inferred mean antibody level half-lives with symbols as in panel a. c) Example of serological measurements and modelled antibody trajectories for a random set of participants (ID-1 to ID-8). Measurements were available before and/or after vaccination (colours) and were either below or above the Roche Elecsys anti-S upper quantification limit of 2960 IU/mL. Modelled trajectories are given in terms of the mean (line) and 95% CrI (shaded area).

### Survival analysis during the Omicron exposure period

In this second part of the analysis, we used survival analysis to evaluate the relationship between the projected anti-S binding antibody levels, as described above, and the hazard of infection during the Omicron exposure period (Figure 1). Data on virologically-confirmed infections during the exposure period (positive test or self-reported negative tests only, see Methods) were available for 967 out of the 1’083 participants, of whom we retained 900 with latest serology after April 1^st^ 2021 (Supplementary Figure S1). The subsample included in this survival analysis was composed of 55% of female participants; 92% were younger than 65 years; 80% had received at least one vaccine dose prior to the start of the Omicron exposure period (December 25^th^ 2021); and 90% had at least one SARS-CoV-2 infection prior to the start of the exposure period (Supplementary Table S3). Out of these 900 participants, 227 had a virologically-confirmed infection during the Omicron exposure period.

We found that the hazard of having an Omicron BA.1/BA.2 infection for individuals with anti-S binding antibody levels higher than a given arbitrary threshold, compared to those with levels below that threshold, decreased down to a three-fold reduction in hazard at a threshold of 800 IU/mL (hazard ratio, HR 0.30, 95% CI 0.22-0.41), and then plateaued for higher antibody level thresholds (Figure 4a). In sensitivity analyses we found consistent effect sizes across antibody thresholds using logistic regression, as well as using different quantiles of the predicted antibody trajectories (Supplementary Material Section S6).

**Figure 4.**
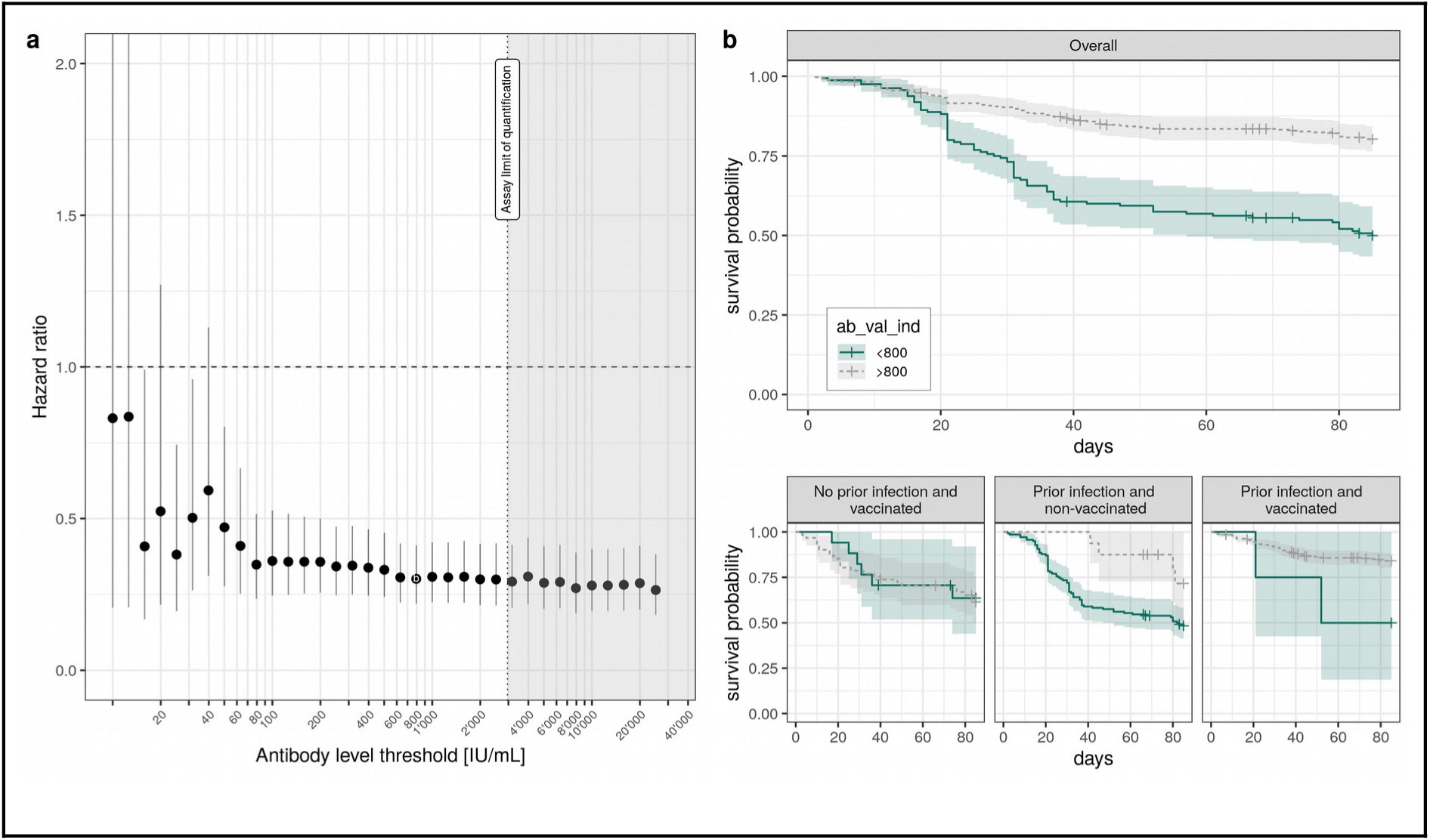
Omicron BA.1/BA.2 infection survival analysis. a) Cox hazard ratio estimates (dots, error bars give the 95% CI) based on proportional hazard models accounting for age and prior infection status across antibody level thresholds. b) Kaplan-Meier curves of the probability of non-infection by SARS-CoV-2 Omicron BA.1/BA.2 stratified by whether predicted antibody levels during the exposure period were above or below 800 IU/mL, shown for the overall analysis dataset, and stratified by infection and vaccination history. Day 0 corresponds to December 25^th^ 2021, when Omicron BA.1 accounted for more than 80% of infections in the state of Geneva (Figure 1c). For this 800 IU/mL threshold, the overall sample size was of N=562 (flowchart in Supplementary Figure S1), subdivided into N=78 for “No prior infection and vaccinated”, N=155 for “Prior infected and non-vaccinated”, and N=329 for “Prior infection and vaccinated”.

We however found that measured antibody-levels do not have the same meaning in terms of correlate of protection whether a participant had a history of infection or not, independently of antibody level (Figure 4b). Similar proportions of Omicron infections were observed among vaccinated participants with no history of infection (and anti-N negative serology) irrespective of their anti-S antibody levels being below or above the 800 IU/mL threshold. Conversely, participants with a history of infection had lower hazard of infection when having antibody levels above the 800 IU/mL threshold, regardless of their vaccination status (Figure 4b, bottom row). Thus, for this anti-S antibody levels threshold, effect estimates stratified by infection and vaccination history showed no significant difference in hazard for uninfected (vaccinated) participants (HR 1.05, 0.36-3.05), and a consistent hazard reduction for participants with a history of infection whether they were vaccinated (HR 0.30, 0.07-1.21) or not (HR 0.45, 0.19-1.06), although small sample sizes in some of these categories yielded wide confidence intervals.

## Discussion

This longitudinal antibody study with follow-up times up to 20 months provided the opportunity to understand long-term anti-SARS-CoV-2 antibody dynamics and to evaluate binding antibody levels from a commercial widely available immunoassay as a correlate of protection against infections during the Omicron BA.1/BA.2 era. Anti-S antibodies persisted up to 20 months after the probable date of infection, with decay dynamics determined by infection and vaccination history. Strongest and longest-lasting antibody boosts occurred with vaccine doses following prior infection. Modelled antibody trajectories enabled the evaluation of binding antibody levels as a correlate of protection against Omicron BA.1/BA.2 infections, for which we found an overall three-fold reduction in the hazard of reporting a positive test for antibody levels above 800 IU/mL. Hazard reduction was however not observed for non-infected participants, indicating that the validity of anti-S binding antibody levels as correlates of protection for Omicron BA.1/BA.2 depends on infection history.

This study extends our previous work showing that anti-SARS-CoV-2 spike antibodies remain detectable after 22 months past probable infection as measured with the Roche anti-S immunoassay (Perez-Saez et al., 2021). Our kinetic modeling results support previous findings indicating that antibody boost is strongest and longest lasting in vaccinees with a history of infection (Dimeglio et al., 2022a; Eyran et al., 2022; Luo et al., 2021). In contrast with previous findings, we found no significant difference in antibody boosting between age groups and slower decay rates in adults 65 years and older (Gallais et al., 2021; Vanshylla et al., 2021). The slower decay rates may be due to age-specific differences in disease severity that we did not account for in these models, thus limiting the comparability of these findings with previous studies due to differences in disease severity profiles. Furthermore, we had a small number of participants over 65 years of age in our study and these age-stratified results should be interpreted with caution. Finally, our results highlight the strong individual-level variability in antibody dynamics, which has been shown in previous antibody kinetic studies (Pelleau et al., 2021; Vanshylla et al., 2021).

Survival analysis results on Omicron BA.1/BA.2 infections are in line with previous findings from vaccine trials targeting the ancestral strain and the Alpha variant, showing that binding antibody levels are an informative correlate of protection against SARS-CoV-2 infection (Feng et al., 2021; Gilbert et al., 2022). These trials had found similar effect sizes of around five-fold reduction in risk of Alpha infections at anti-S antibody levels of 600 IU/mL (Feng et al., 2021), and a halving of hazard by 10-fold increase in anti-S titers for ancestral strain infections (Gilbert et al., 2022). Moreover our results are in line with available studies on Omicron BA.1/BA.2 subvariants which have also found binding antibody levels to be correlates of protection against infection using in-house immunoassays (Hertz et al., 2022; Zar et al., 2022). On the other hand, we did not find differences in the hazard of having an Omicron BA.1/BA.2 infection with anti-S antibody levels below or above a certain threshold in the non-infected vaccinated group, as opposed to results reported for Delta infections (Wei et al., 2021). Notably, this finding is supported by our recent work on neutralization capacity in the Geneva population (Zaballa et al., 2022). Using the same immunoassay as in this study and a cell-free Spike trimer-ACE2 binding-based surrogate neutralization assay (Fenwick et al., 2021), we did not observe any significant correlation between anti-S binding and neutralizing antibody levels against Omicron subvariants in uninfected participants, as opposed to previously infected participants (Zaballa et al., 2022). These results can be linked to growing evidence that hybrid immunity (infection plus vaccination) provides the strongest protection against Omicron subvariant infections (Altarawneh et al. 2022, Zar et al. 2022, Golddblatt 2022). This infection history-specificity thus warrants care in the interpretation of binding antibodies as correlates of protection against Omicron sub-lineages, and could be immunoassay-dependent.

We note that it remains unclear whether these correlate of protection results extend to subsequent Omicron subvariants (BA.4, BA.5, BA.2.75, BQ.1 and othres), which have been found, thanks to specific mutations, to have stronger immune-evasion capacity than the parent BA.1 strain (Cao et al., 2022; Tan et al., 2022). Changes in immune evasion capacity may theoretically, if multiple mutations accumulate on the spike protein, impact the level of binding antibodies at which hazard reduction occurs as well as its effect size. Moreover, our longitudinal serology follow-up was conducted before the circulation of the Omicron lineage in Geneva. The interpretation of anti-S antibody levels measured with this immunoassay following Omicron infections might need to be revisited in the light of evidence of reduced test sensitivity towards antibodies targeting the Omicron Spike protein (Springer et al., 2022).

This study has several limitations. Firstly, we only used the Roche Elecsys assay, which measures total anti-S antibodies (IgA/M/G), whose levels may correlate differently with overall immune function following infection or vaccination; other immunoassays may have different antibody binding characteristics. Secondly, analyses in the 65+ subgroup are limited by the small number of participants. Thirdly, our survival analysis to assess correlates of protection was based on modeled antibody trajectories, and not on measurements at defined time points as done in studies available from vaccine trails (Gilbert et al., 2022). Although modeled trajectories matched well antibody participant-level measurements, the survival analysis results are subject to modeling uncertainty. While sensitivity analysis using the 2.5% and 97.5% prediction quantiles yielded qualitatively similar correlate of protection results, other sources of modeling uncertainty cannot be excluded. Fourthly, a large proportion of virologically-confirmed infections (44%, 101/227) were self-reported as opposed to the other 56%, which were directly extracted from the state COVID-19 test registry (ARGOS). Reassuringly, of the 1’083 participants in our longitudinal sample for whom tests in the registry were available, self-reported positive tests with matching dates were reported in 82% (491/599) of cases, thus suggesting a reasonable sensitivity of self-reporting. Finally, both Omicron BA.1 and BA.2 subvariants circulated in the canton of Geneva during the study exposure period and sequencing information on infection was not available, thus precluding a differential correlate of protection analysis for both subvariants.

Overall, this study extends findings against previous SARS-CoV-2 variants showing that anti-S binding antibody levels are a valid correlate of protection against Omicron BA.1/BA.2 infections. Importantly, we found that the validity of antibody levels as correlate of protection depends on infection history as quantified with the immunoassay used in this study. Our results highlight the imperfect nature of protection after vaccination and/or infection. Even with perfect knowledge of infection and vaccination histories, inference about population-level immunity continues to pose challenges. Future studies may benefit from the modeling framework developed in this study to leverage longitudinal measurements to epidemiological outcomes. Taken together, these conclusions motivate further investigation of how immune landscape and immunoassay characteristics determine the interpretation of serological surveys into population-levels of protection to inform public health decisions.

## Data Availability

Data produced in this study is accessible to researchers upon reasonable request.

https://github.com/UEP-HUG/serosuivi_2021_public

## Acknowledgements

We thank Aglaé Tardin and the team of General Directorate of Health of Geneva for giving us access to the ARGOS state registry, and the Hôpital de La Tour and the Clinique de Carouge for allowing us to use their premises for the recruitment of participants. We are deeply grateful to all the participants, without whom this study would not have been possible.

