## Supplementray Material for "Long term anti-SARS-CoV-2 antibody kinetics and correlate of protection against Omicron BA.1/BA.2 infection"

#### Contents

**S1 Study sample** **S2**
S1.1 Definition of prior infection status . . . . . S2
S1.2 Survival analysis exclusion criteria . . . . . S2

**S2 Immunoassay quality controls** **S3**

**S3 Additional antibody trajectory data** **S4**
S3.1 Anti-S . . . . . S4
S3.2 Anti-N . . . . . S5

**S4 Antibody kinetics model** **S5**
S4.1 Base model . . . . . S5
S4.2 Multi-boosting model . . . . . S6
S4.3 Unknwon infection dates . . . . . S6
S4.4 Priors . . . . . S7
S4.5 Parameter estimates . . . . . S8

**S5 Survival analysis details** **S9**

**S6 Survival sensitivity analysis** **S11**

### S1 Study sample

The flowchart of participants in the study is presented in Figure S1. The full event dataset of retained participants is shown in Figure S2.

#### S1.1 Definition of prior infection status

Both the antibody kinetic model and the survival analysis required determining participants' status of prior SARS-CoV-2 infection. We defined prior infection as the at least one of the following conditions prior to the date of interest: (a) a positive test results from the state registry (ARGOS database), (b) a self-reported test not followed by a negative anti-N result within 3 months of the self-reported test date to account for positive false-positive reporting, or (c) a positive anti-N serology.

#### S1.2 Survival analysis exclusion criteria

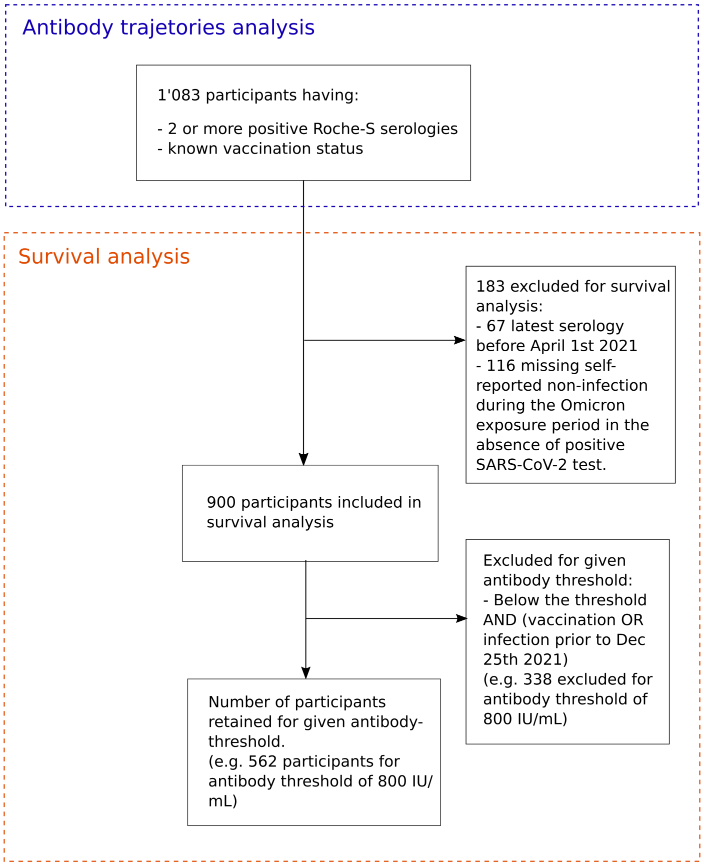


Figure S1: Flowchart of study participants. Participants in this study were all selected from the Specchio-COVID19 longitudinal cohort in the state of Geneva, Switzerland. Among selected participants for antibody kinetics analysis we further excluded participants according to infection status information for the survival analysis.

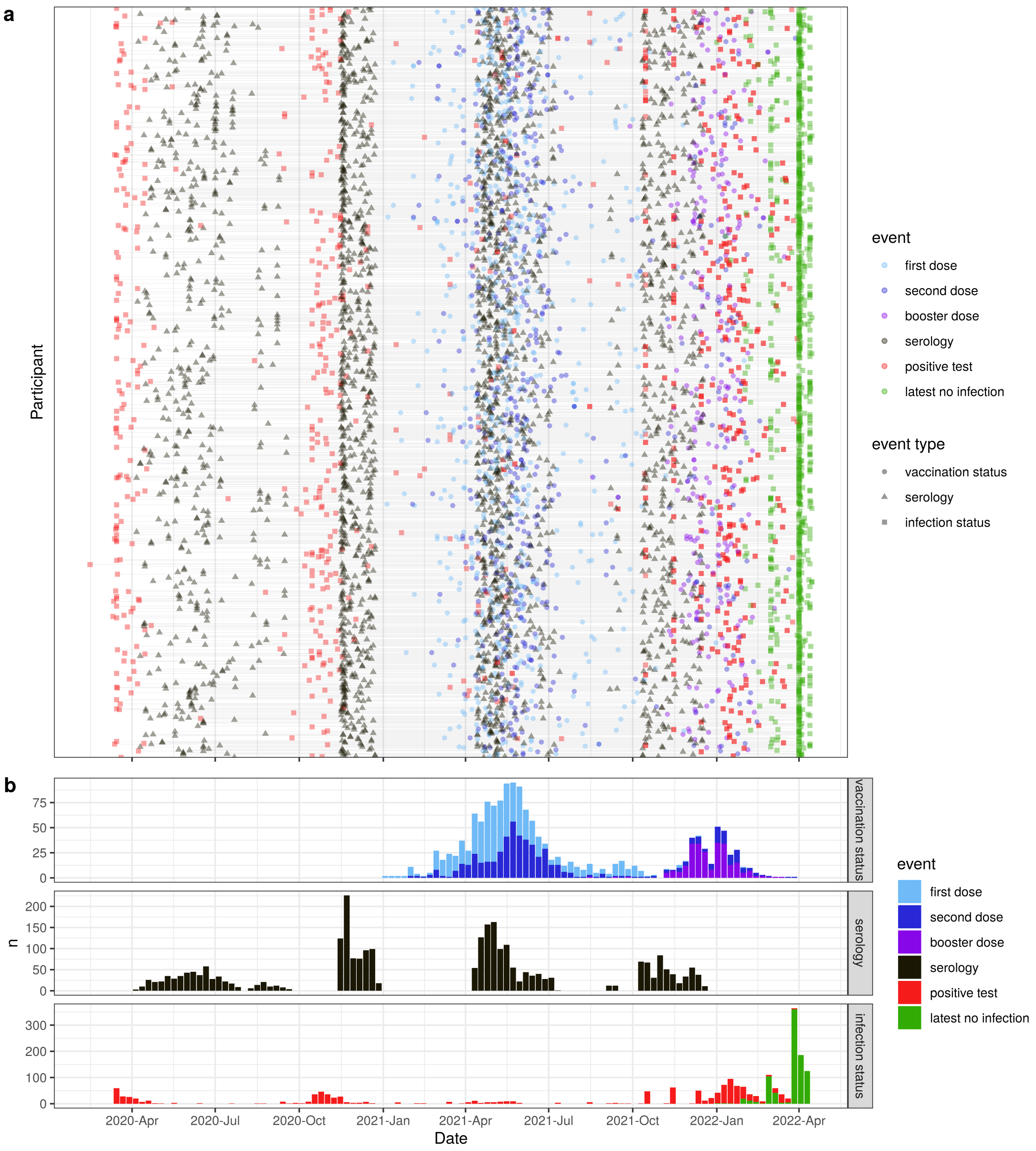


Figure S2: Participant-level events for all participants in the study. a) Timeline of events. Each horizontal line represents one participant, and points represent different types of events (serology, vaccination, tests). b) Weekly counts of distinct events, stratified by event type.

### S2 Immunoassay quality controls

Table S1 shows the means, standard deviations and coefficient of variations for the external quality controls used to assess inter-lot variability of the Roche anti-S and anti-N immunoassays as described in Perez-Saez et al. (2021).

Table S1: Quality controls of the Roche Elecsys anti-N (Roche-N, in arbitray units) and Roche Elecsys anti-spike (Roche-S, in U/mL) immunoassays. CQI lot IDs refer to different lots of external positive controls used to compute coefficients of variation.

| CQI lot ID | mean | sd | CV [%] |
|---|---|---|---|
| **Roche-N** | | | |
| ID-1 | 4.0 | 0.3 | 7.3 |
| ID-2 | 10.9 | 0.6 | 5.2 |
| ID-3 | 6.0 | 0.2 | 4.1 |
| **Roche-S** | | | |
| ID-1 | 5.0 | 0.4 | 7.3 |
| ID-2 | 24.9 | 1.5 | 6.2 |
| ID-3 | 10.7 | 0.3 | 2.6 |

### S3 Additional antibody trajectory data

#### S3.1 Anti-S

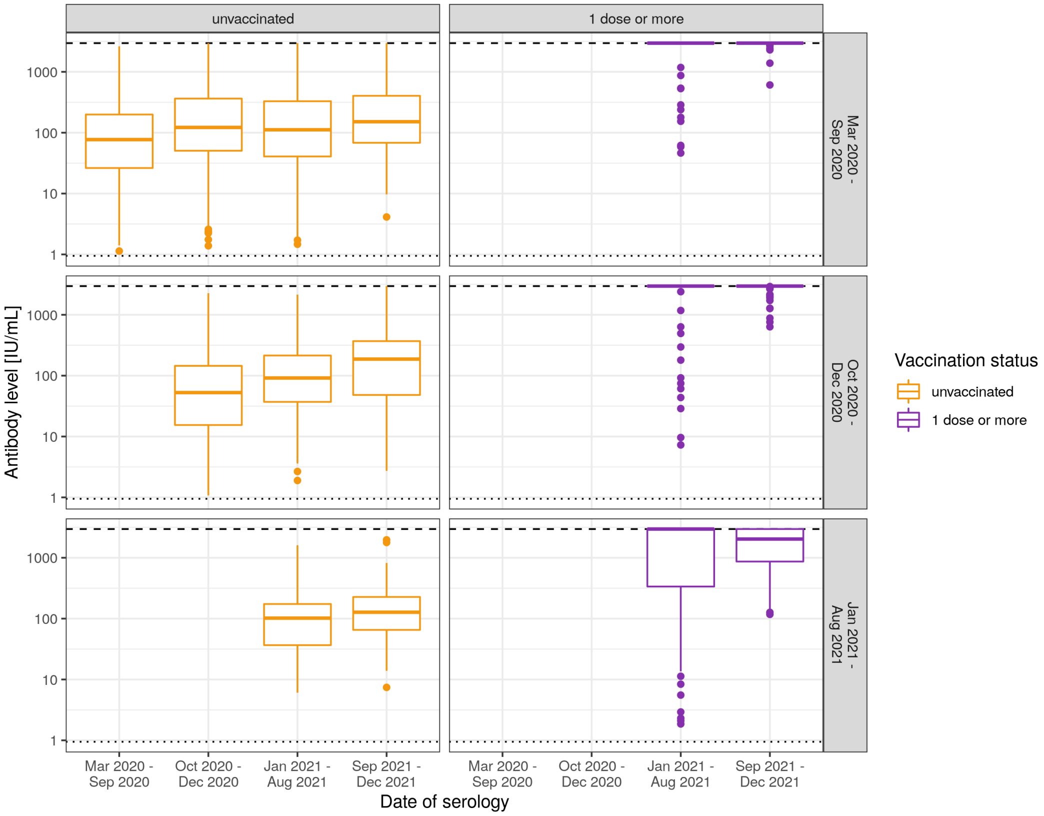


Figure S3: Distribution of anti-S antibody levels by visit date. Boxplots are stratified by vaccination status at time of serology (columns) and period of first serology (rows).

#### S3.2 Anti-N

Dynamics in anti-N antibody levels as quantified by the Roche immunoassay are shown in Figure S4.

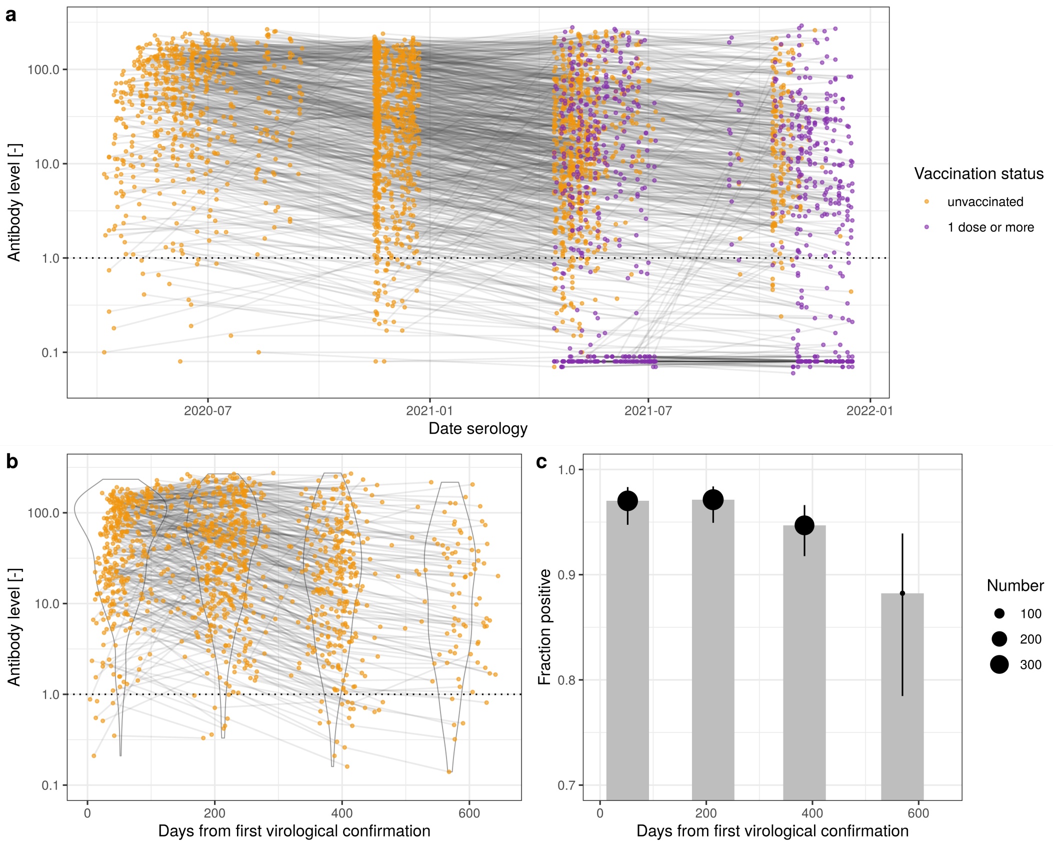


Figure S4: Anti-N antibody trajectories. a) Trajectories for all study participants, colored by vaccination status. b) Antibody level trajectories in time from virological confirmation for participants with at least one anti-N positive serology and known date of positive test (N=478), along with violin plots of antibody level in discrete categories of time post confirmation (0-149, 150-249, 250-449, 450+ days). Threshold for positivity, as indicated by the manufacturer, is index larger or equal to 1.0 and indicated with horizontal dotted line. c) Fraction of participants with at least one anti-N positive serology and known date of positive test (N=478) above positivity threshold with time after first virological confirmation for time categories as in (b), along with 95% CIs.

### S4 Antibody kinetics model

#### S4.1 Base model

We used an antibody kinetics model to infer trajectories of antibody levels for each participant as developed for SARS-CoV-2 (Pelleau et al. 2021). The bi-phasic model we built on accounted for the antibody trajectory for participant $i$ after an initial boost through infection or vaccinations through the contribution of short-lived and long-lived antibody-secreting cells as:

$$y_i(t) = \alpha + \beta_i \times \left( \rho_i \frac{e^{-\gamma_{s_i}(t-\delta_i)} - e^{-\nu_i(t-\delta_i)}}{\nu_i - \gamma_{s_i}} + (1-\rho_i) \frac{e^{-\gamma_{l_i}(t-\delta_i)} - e^{-\nu_i(t-\delta_i)}}{\nu_i - \gamma_{l_i}} \right) \quad (1)$$

where $\alpha$ is the baseline level, $\beta_i$ is the antibody boost, $\rho_i$ is the proportion of short-lived vs. long-lived antibody-secreting cells, and $\gamma_{s_i}$, $\gamma_{l_i}$ and $\nu_i$ are the rates of short-live, long-lived and antibody decay rates, and $\delta_i$ is the delay between exposure and humoral response. All parameters depend on the characteristics of the participant (eg. age, sex) and the boost (eg. infection, re-infection, vaccine dose), encoded in covariate matrix $\boldsymbol{X}$, accounting for participant-level heterogeneity:

$$\begin{aligned}
\beta_i &= \exp(\boldsymbol{b}_\beta \boldsymbol{X}_i + u_{\beta_i}), \\
\gamma_{s_i} &= \exp(\boldsymbol{b}_{\gamma_s} \boldsymbol{X}_i + u_{\gamma_{s_i}}), \\
\gamma_{l_i} &= \exp(\boldsymbol{b}_{\gamma_l} \boldsymbol{X}_i + u_{\gamma_{l_i}}), \\
\nu_i &= \exp(\boldsymbol{b}_\nu \boldsymbol{X}_i + u_{\nu,i}), \\
\rho_i &= logit^{-1}(\boldsymbol{b}_\rho \boldsymbol{X}_i), \\
u_{\beta_i} &\sim \mathcal{N}(0, \sigma_\beta), \\
u_{\gamma_{s_i}} &\sim \mathcal{N}(0, \sigma_{\gamma_s}), \\
u_{\gamma_{l_i}} &\sim \mathcal{N}(0, \sigma_{\gamma_l}), \\
u_{\nu,i} &\sim \mathcal{N}(0, \sigma_\nu),
\end{aligned}$$

where $u$ are the participant-level random effects with distinct variances.

The likelihood for a given observation $y_i^{obs}$ was assumed to follow a normal distribution with known coefficient of variation $cv = 7.3\%$ based on the maximum CV in Table S1, which can be expressed through log-transformation as:

$$\mathcal{L}(y_i) = \mathcal{N}(log(y_i^{obs})|log(y_i), cv).$$

#### S4.2 Multi-boosting model

Participants in this study may have experienced multiple antibody boosts from infection and/or vaccination origin. We assume that successive antibody boosts have an additive effect on the total antibody level:

$$y_i(t) = \alpha + \sum_{b=1}^{B_i} \beta_{i,b} \times \left( \rho_{i,b} \frac{e^{-\gamma_{s_{i,b}}(t-\tau_{i,b}-\delta_{i,b})} - e^{-\nu_{i,b}(t-\tau_{i,b}-\delta_{i,b})}}{\nu_{i,b} - \gamma_{s_{i,b}}} + (1-\rho_{i,b}) \frac{e^{-\gamma_{l_{i,b}}(t-\tau_{i,b}-\delta_{i,b})} - e^{-\nu_{i,b}(t-\tau_{i,b}-\delta_{i,b})}}{\nu_{i,b} - \gamma_{l_{i,b}}} \right),$$

where $B_i$ is the number of antibody boosts of participant $i$, and $\tau_{i,b}$ is participant's relative time from initial boost to boost $b$.

#### S4.3 Unknwon infection dates

In eq.1 the infection date is assumed to be known. For participants for which we have a positive test we assume that the date of infection is the same as the date of positive test. However dates of positive test were unreported for about half of participants with history of infection based on anti-N serology (Table 1 in main text). We therefore marginalize over possible delays between infection and serological visits:

$$\mathbb{P}(y_i|\mathbf{\Theta}) = \sum_{\tau_i'=0}^{\tau'_{max}} \mathcal{N}(log(y_i^{obs})|log(y_{i,\tau_i'}), cv) \times \mathbb{P}(\tau_i'),$$

$$y_{i,\tau_i'} = \alpha + \beta_i \times \left( \rho_i \frac{e^{-\gamma_{s_i}(t-\tau_i'-\delta_i)} - e^{-\nu_i(t-\tau_i'-\delta_{i,b})}}{\nu_i - \gamma_{s_{i,b}}} + (1-\rho_{i,b}) \frac{e^{-\gamma_{l_{i,b}}(t-\tau_{i,b}'-\delta_i)} - e^{-\nu_{i,b}(t-\tau_i'-\delta_i)}}{\nu_{i,b} - \gamma_{l_i}} \right),$$

$$\mathbb{P}(\tau_i') = \mathbb{P}(t_i^s = t|t_i^v) = \frac{cases_{t+\delta}}{\sum_{t\prime=t_0}^{t_i^v} cases_{t'+\delta}},$$

where $\mathbf{\Theta}$ is the set of model parameters, $\mathbb{P}(\tau_i')$ is the probability of participant $i$ having been infected on day $\tau_i$, $t_i^v$ is the date of the serological visit, and $\tau_i^{max}$ is the furthest possible infection date given the start of the pandemic at $t_0$ and previous serologies. We assume that the conditional probability of infection on a given date is proportional to the number of reported cases relative to the total reported cases in the allowed time range for participant $i$ (from the most recent serology date with a positive anti-N serology to the one with a negative anti-N serology, or to the start of the pandemic if no anti-N serologies are available prior to the first positive one). Following Perez-Saez et al. (2021), we correct for differences in case reporting delays and reporting probability using reporting delays and case-to-infection ratios for the state of Geneva reported in Stringhini, Wisniak, et al. (2020) and Stringhini, Zaballa, et al. (2021). For computational reasons and to mitigate issue with reporting delays, we compute the probability of infection for mutually-exclusive 10-day periods instead of at a daily time step.

#### S4.4 Priors

We set the following priors for model parameters:

$$\begin{aligned}
\alpha &\sim \mathcal{N}(0.1, 1), \\
\delta &\sim \mathcal{N}(5, 2), \\
exp(\boldsymbol{b_\beta}[1]) &\sim \mathcal{N}(150, 50), \\
t_{\gamma_s}^{\frac{1}{2}} = \log(2)/exp(\boldsymbol{b_{\gamma_s}}[1]) &\sim \mathcal{N}(10, 3), \\
t_{\gamma_l}^{\frac{1}{2}} = \log(2)/exp(b_{\boldsymbol{\gamma_l}}[1]) &\sim \mathcal{N}(500, 100), \\
t_{\nu}^{\frac{1}{2}} = \log(2)/exp(\boldsymbol{b_\nu}[1]) &\sim \mathcal{N}(60, 10), \\
logit^{-1}(\boldsymbol{b_\rho}[1]) &\sim beta(5, 1), \\
\boldsymbol{b_{\gamma_s}}[2:K] &\sim \mathcal{N}(0, .5), \\
\boldsymbol{b_{\gamma_l}}[2:K] &\sim \mathcal{N}(0, .5), \\
\boldsymbol{b_\nu}[2:K] &\sim \mathcal{N}(0, .5), \\
\boldsymbol{b_\rho}[2:K] &\sim \mathcal{N}(0, .5), \\
\sigma_\beta &\sim \mathcal{N}(0, 2), \\
\sigma_{\gamma_s} &\sim \mathcal{N}(0, 2), \\
\sigma_{\gamma_l} &\sim \mathcal{N}(0, 2), \\
\sigma_\nu &\sim \mathcal{N}(0, 2),
\end{aligned}$$

where [1] denotes the first element of parameter vectors corresponding to the intercept, and $[2:K]$ denotes the values corresponding to other covariate combinations. We set priors on the half-lives $t_{\gamma_s,\gamma_l,\nu}^{\frac{1}{2}}$ of short-lived, long lived cells and antibodies respectively instead of regression parameters for ease of interpretation based on previous results for SARS-CoV-2 antibody kinetics (Pelleau et al. 2021). Similarly we set a beta prior on the inverse-logit of the regression parameters controlling the proportion of short-lived vs. long-lived cells $\boldsymbol{b_\rho}$. The baseline amount of antibodies $\alpha$ and boosting level $exp(b_\beta)$ is given in international antibody level units and delay parameter $\delta$ and half-lies in days.

Table S2: Kinetic model parameter estimates. Estimates are given by age category for the mean boost level and the mean antibody half-life in terms of the mean and the 95% credible interval. Estimates were rounded to the nearest tens.

| Infection/vaccination | Boost level [IU/mL] | Antibody half-life [days] |
|---|---|---|
| **Age: 18-64** | | |
| dose1 | 720 (300-1’480) | 170 (60-360) |
| dose1+dose2 | 4’700 (2’160-8’860) | 50 (30-100) |
| dose1+dose2+booster | 430 (140-1’050) | 170 (60-410) |
| infected | 370 (270-460) | 150 (130-160) |
| infected+dose1 | 12’220 (6’300-21’630) | 140 (70-250) |
| infected+dose1+dose2 | 1’030 (370-2’240) | 190 (70-430) |
| infected+dose1+dose2+booster | 470 (160-1’120) | 170 (60-400) |
| infected+infected2 | 280 (100-610) | 100 (40-240) |
| infected+infected2+dose1 | 2’360 (830-5’030) | 250 (90-540) |
| infected+infected2+dose1+dose2 | 530 (150-1’370) | 170 (60-420) |
| infected+infected2+dose1+dose2+booster | 420 (130-1’020) | 170 (50-390) |
| **Age: 65+** | | |
| dose1 | 910 (300-2’200) | 350 (100-910) |
| dose1+dose2 | 5’910 (2’080-13’880) | 110 (40-250) |
| dose1+dose2+booster | 530 (130-1’460) | 350 (90-960) |
| infected | 460 (210-870) | 310 (150-560) |
| infected+dose1 | 15’370 (5’740-35’290) | 280 (100-630) |
| infected+dose1+dose2 | 1’300 (360-3’400) | 390 (110-1020) |
| infected+dose1+dose2+booster | 590 (160-1’570) | 350 (90-940) |
| infected+infected2 | 360 (100-930) | 210 (60-560) |
| infected+infected2+dose1 | 2’970 (820-7’720) | 510 (140-1360) |
| infected+infected2+dose1+dose2 | 670 (160-1’900) | 360 (90-1000) |
| infected+infected2+dose1+dose2+booster | 530 (130-1’420) | 350 (90-950) |

#### S4.5 Parameter estimates

Table S2 shows the parameter estimates of boost level and antibody half-life.

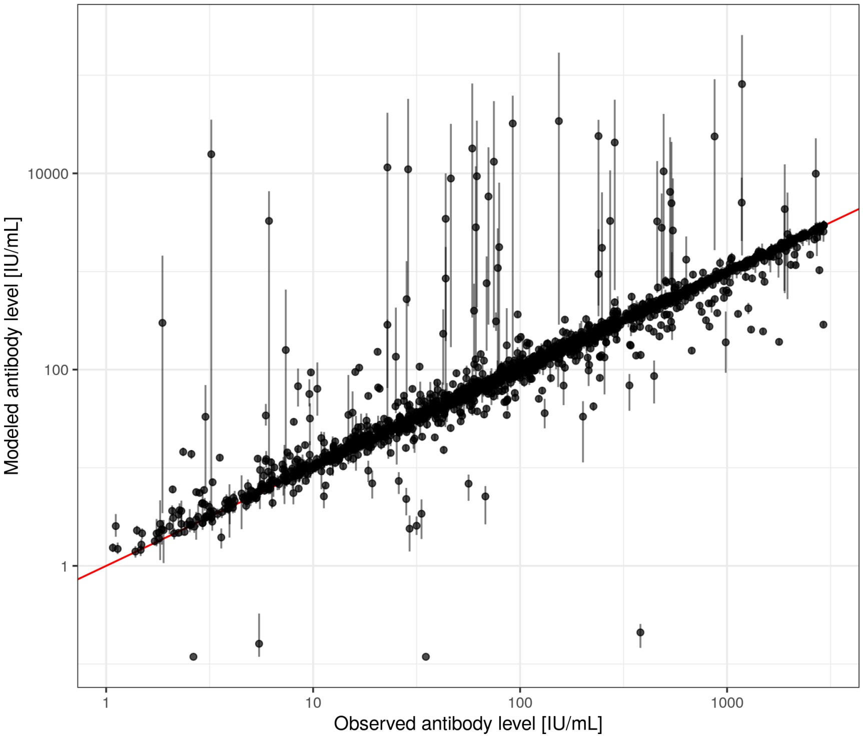


Figure S5: Comparison of observed and modeled antibody levels. Modeled levels are given in terms of the mean (dots) and 90% credible intervals (bars).

### S5 Survival analysis details

We model the impact of antibody levels through a Cox proportional hazards model to test the assumption that antibody levels have an effect on the hazard of reporting a positive Omicron test.

Our causal assumptions on the link between factors involved in a positive test during the Omicron infection period are detailed the directed acyclical graph (DAG) in Figure S6. We assume that unobserved covariates influence participant exposure to infection, and that age affects both infection and vaccination. In turn, prior infection and vaccination affect the level of antibodies in each participant. We assume that vaccination status does not influence the probability of reporting a positive test through other pathways than through antibody levels.

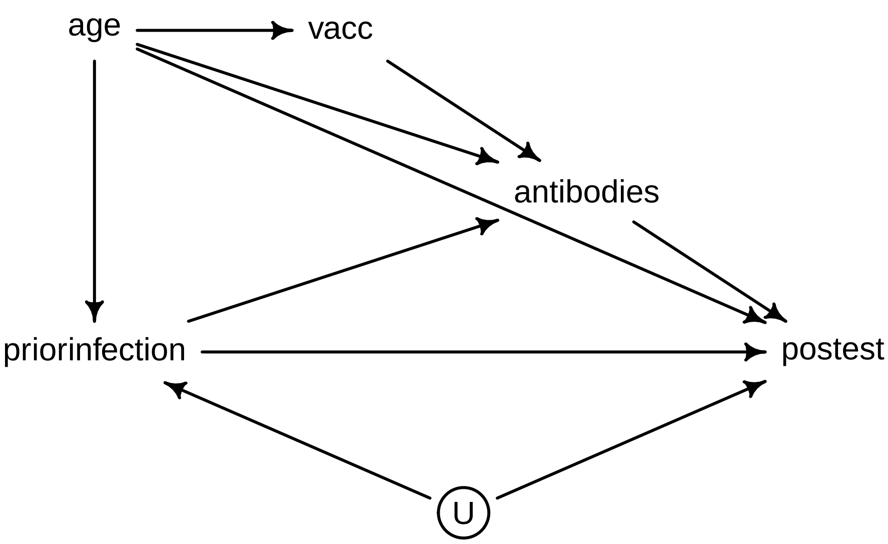


Figure S6: DAG of effect of antibody levels on reporting a positive Omicron test. antibodies: whether antibody level is above or below given threshold, postest: positive virological test during Omicron exposure period, U: unobserved factor controlling probability of infection, priorinfection: history of SARS-CoV-2 infection, age: age category, vacc: vaccinated or unvaccinated.

The hazard rate for reporting a positive test is then:

$$\lambda_i(t) = \lambda_0(t)\exp(\beta_0 antibodies + \beta_1 age + \beta_2 priorinfection).$$

The characteristics of participants retained in the survival analysis are given in Table S3.

Table S3: Characteristics of participants retained for survival analysis during the period of Omicron exposure.

| Characteristic | N = 900[1] |
|---|---|
| infected_prior | 811 (90%) |
| vaccinated | 716 (80%) |
| sex | |
| female | 498 (55%) |
| male | 402 (45%) |
| age_cat | |
| [18,65) | 827 (92%) |
| [65,Inf] | 73 (8.1%) |

[1]n (%)

#### S6 Survival sensitivity analysis

Figure S7 shows the comparison of effect estimates between the logistic and Cox proportional hazard models. Figure S8 shows the effect estimates when using the 2.5% and 97.5% posterior quantiles of modeled antibody trajectories instead of the mean.

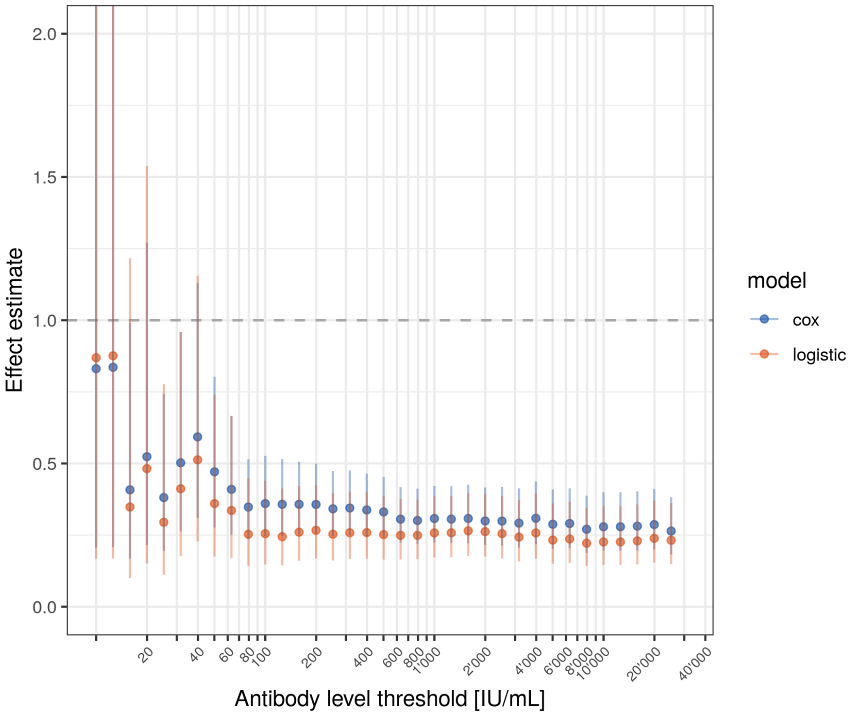


Figure S7: Comparison of effect estimates between logistic and Cox proportional hazard models of being above a given antibody level threshold.

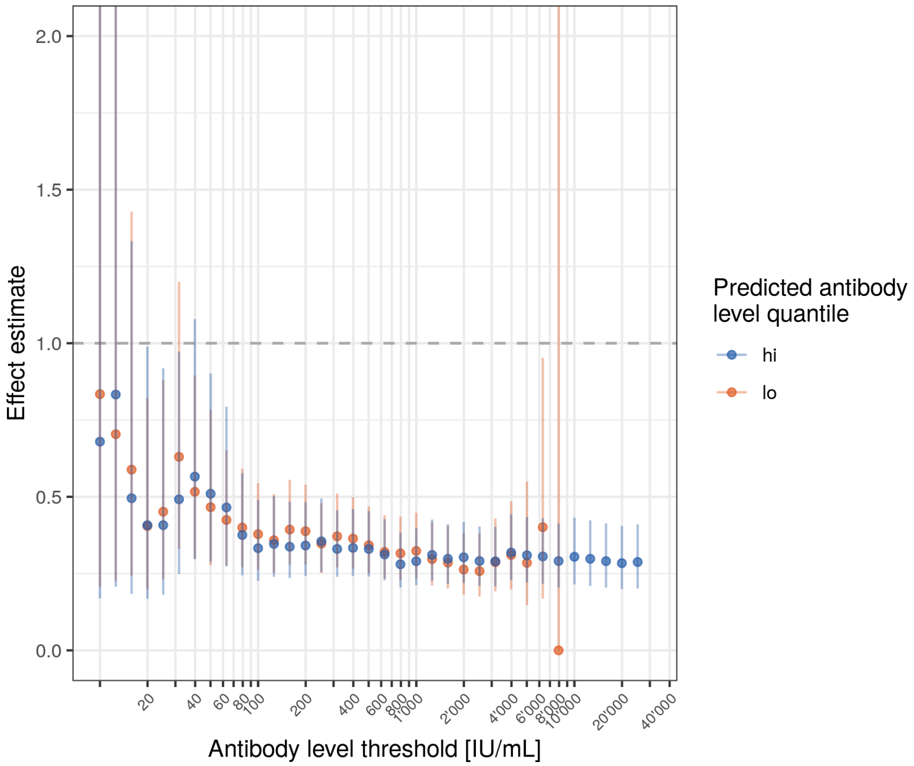


Figure S8: Comparison of effect estimates when using the 2.5% (lo) and 97.5% (hi) credible intervals of modeled antibody trajectories.
